# How the clinical research community responded to the COVID-19 pandemic: An analysis of the COVID-19 clinical studies in ClinicalTrials.gov

**DOI:** 10.1101/2020.09.16.20195552

**Authors:** Zhe He, Arslan Erdengasileng, Xiao Luo, Aiwen Xing, Neil Charness, Jiang Bian

## Abstract

**Objective:** The novel coronavirus disease (COVID-19), broke out in December 2019, and is now a global pandemic. In the past few months, a large number of clinical studies have been initiated worldwide to find effective therapeutics, vaccines, and preventive strategies for COVID-19. In this study, we aim to understand the landscape of COVID-19 clinical research and identify the gaps such as the lack of population representativeness and issues that may cause recruitment difficulty.

**Materials and Methods:** We analyzed 3,765 COVID-19 studies registered in the largest public registry - ClinicalTrials.gov, leveraging natural language processing and using descriptive, association, and clustering analyses. We first characterized COVID-19 studies by study features such as phase and tested intervention. We then took a deep dive and analyzed their eligibility criteria to understand whether these studies: (1) considered the reported underlying health conditions that may lead to severe illnesses, and (2) excluded older adults, either explicitly or implicitly, which may reduce the generalizability of these studies to the older adults population.

**Results:** Most trials did not have an upper age limit and did not exclude patients with common chronic conditions such as hypertension and diabetes that are more prevalent in older adults. However, known risk factors that may lead to severe illnesses have not been adequately considered.

**Conclusions:** A careful examination of existing COVID-19 studies can inform future COVID-19 trial design towards balanced internal validity and generalizability.

## Introduction

The severe acute respiratory syndrome coronavirus 2 (SARS-CoV-2) and the associated coronavirus disease (COVID-19) broke out in December 2019 and has quickly become a global pandemic with serious health and social consequences [1]. As of October 27, 2020, more than 70 million confirmed cases have been reported around the world and about one-fifth are from the U.S. [2]. Globally, more than 1,592,000 people have died due to COVID-19 and 295,000 in the U.S. alone. In April 2020, the National Institutes of Health (NIH) launched the Accelerating COVID-19 Therapeutic Interventions and Vaccines (ACTIV) public-private partnership to prioritize and speed up the clinical evaluation of the most promising treatments and vaccines [3]. In July 2020, NIH released its strategic plan for COVID-19 research to speed up the development of treatments, vaccines, and diagnostics [4]. Traditionally, it may take years to discover, develop, and evaluate a therapeutic agent; nevertheless, for COVID-19, the goal has been to compress the timeline to months while continuing to apply rigorous standards to ensure safety and efficacy. Strategies such as applying complex computer-generated models of SARS-Cov-2 and its biological processes to determine key interactions and pathways have been applied to develop therapeutic agents and vaccines (e.g., monoclonal antibodies) to neutralize the virus. A significant efforts have also been made to screen existing drugs approved for other indications to treat COVID-19 [4]. Recently, the COVID-19 vaccine developed and manufactured by Pfizer-BioNTech has been approved to be administered in a few countries including the United Kingdom [5], Canada [6], and the US [7].

Clinical studies, especially randomized controlled trials (RCT), are the gold standard for evaluating the efficacy and safety of a treatment. Regardless of the techniques (e.g., in vivo, in silico, or in vitro) used for drug discovery, the therapeutics and vaccines have to go through three phases of clinical trials to evaluate their efficacy and safety before approvals (e.g., by the U.S. Food and Drug Administration [FDA]) can be granted for mass production and use in the general population. In the past few months, many COVID-19 clinical studies have been launched around the world, leading to situations where studies have to compete for participants from the same pool of eligible participants. Trials such as those for the promising drug – *Remdesivir* – were suspended due to the lack of trial participants in China [8]. Other issues such as population representation are also critical. In the past, older adults are often excluded from clinical trials with overly restrictive exclusion criteria, which lead to concerns on the generalizability of those clinical studies across many disease domains [9]. A recent New York Times article conjectured that older adults are left out form COVID-19 trials [10]. It is therefore important to understand the landscape of COVID-19 clinical research and further identify the gaps and issues that may cause delays in patient recruitment and the lack of real-world population representativeness, especially for older adults.

To date, 12 other studies have analyzed registered COVID-19 clinical studies [11-22] (see Supplementary Material I for details). For example, Wang et al. analyzed the basic characteristics and the drug interventions of 306 COVID-19 trials from ClinicalTrials.gov as of April 3, 2020 [11]. Paudi et al. analyzed 1,551 COVID-19 studies registered between March 1, 2020 and May 19, 2020 in ClinicalTrials.gov and focused on basic characteristics, primary and secondary outcomes, and study design [12]. Kim et al. evaluated the impact of frequently used quantitative eligibility criteria in 288 COVID-19 studies (as of June 18, 2020) on the recruitment and clinical outcomes using the EHR data of COVID-19 patients in Columbia University Irving Medical Center [22]. While most of these studies analyzed the basic characteristics of the included trials, some studies further analyzed the interventions [18-21], locations [13,14,18], data monitoring characteristics [14], timing of the registration and enrollment [15,21], outcomes [19,20], risk of biases [16], Medical Subject Heading (MeSH) keywords (Words or phrases that best describe the protocol) [19], sample size [17], and a subset of eligibility criteria [16,22]. Nonetheless, existing studies have not comprehensively investigated the qualitative eligibility criteria and the consideration of known risk factors of severe illnesses in COVID-19 clinical studies. To do so, advanced approaches such as natural language processing (NLP) are necessary. As such, we can gain a better understanding of the landscape of COVID-19 research and answer a number of important research questions: (1) What eligibility criteria are used in COVID-19 clinical studies? Are these criteria too restrictive? (2) Further, as more COVID-19 cases have been identified and treated in the past 8 months, we have accumulated important knowledge on the underlying health conditions and other risk factors that may cause severe illness among COVID-19 patients (e.g., hypertension and diabetes) [23]. Have existing clinical studies sufficiently considered these known risk factors? (3) Last but not the least, because of the concerns on study generalizability in older adults, it is of interest to assess whether the COVID-19 clinical studies excluded subjects with common chronic conditions that are prevalent in older adults, potentially leading to their underrepresentation.

In this study, we conducted a systematic analysis of the registered clinical studies on COVID-19 (as of November 27, 2020) from ClinicalTrials.gov to answer the aforementioned research questions. The contribution of this paper is multi-fold: (1) it systematically summarizes various important aspects of the COVID-19 clinical studies; (2) it identifies the research gaps on the risk factors related to serious illness caused by COVID-19; (3) it groups COVID-19 studies based on their eligibility criteria, and (4) it identifies salient exclusion criteria that may implicitly exclude older adults, who are most vulnerable and should be studied when evaluating the efficacy and safety of COVID-19 treatments and vaccines. Our findings could inform future trials designed for COVID-19 treatment and prevention and identify strategies to rapidly but appropriately stand up a large number of clinical studies for future pandemics similar to COVID-19.

## Materials and Methods

### Data Source

ClinicalTrials.gov, built and maintained by the U.S. National Library of Medicine, is the largest clinical study registry in the world [24]. In the U.S., all drugs and devices regulated by the U.S. Food and Drug Administration (FDA) are required to be registered on the ClinicalTrials.gov. ClinicalTrials.gov is thus considered as the most comprehensive trial registry in the world and has been widely used for secondary analysis [25].

### Dataset Acquisition and Processing

From ClinicalTrials.gov, we downloaded records of 4,028 clinical studies that are tagged with a condition of “COVID-19” or “SARS-CoV-2” on November 27, 2020. We excluded studies tagged with the study type “Expanded Access” and studies that were tagged as patient registries, leaving 3,765 records that met our inclusion criteria. We extracted the NCTID (an unique identifier of a study record), conditions, agency, agency class, brief summary, detailed summary, status, start date, eligibility criteria, enrollment, study phase, study type, intervention type, intervention name, study design (i.e., allocation, masking, observation model, time perspective), primary purpose, and endpoint classification. We split the eligibility criteria into inclusion criteria and exclusion criteria and further extracted individual criteria for natural language processing. To identify the top frequently tested drugs, we extracted the drugs information from the “intervention” field from the study record. We used QuickUMLS to normalize the drug names and removed the dosage information before analyzing their frequencies.

### Consideration of Risk Factors in COVID-19 Clinical Studies

We first identified known risk factors of COVID-19 from online resources such as the Centers for Disease Control and Prevention (CDC) [26] and Mayo Clinic [27] (as of July 17, 2020). Then, we coded the risk factors with the concepts from Unified Medical Language System (UMLS). To do so, we used the risk factor terms as the input and identified their corresponding Concept Unique Identifiers (CUIs) using QuickUMLS [28] with the default setting (Jaccard similarity threshold > 0.8, all semantic types included). As a concept of the UMLS is associated with its synonyms from UMLS source ontologies, we were able to unify all the terms mentioned in the text. ***Table 1*** lists the risk factors that may lead to severe COVID-19 and their associated UMLS CUIs. This list was used as a dictionary in QuickUMLS [28] to identify risk factors from the study description. As reported in Soldaini et al. [28], QuickUMLS achieved better performance than MetaMap and cTAKES on a number of benchmark corpora. Nevertheless, we manually reviewed the risk factors extracted by QuickUMLS on a random sample of 100 clinical studies; and QuickUMLS achieved a precision of 91%^1^. We also identified the studies that used these risk factors in the inclusion/exclusion criteria using the parsing results of the eligibility criteria parsing tool [29] described below. T-test and analysis of variance (ANOVA) were employed to assess the association between the number of risk factors in the trial descriptions with the study type, intervention type, and primary purpose.

**Table 1.** UMLS CUIs for the risk factors for severe illness among COVID-19 patients reported by CDC and Mayo Clinic websites

| **Risk Factors** | **UMLS CUIs** |
| --- | --- |
| Old age | C0231337, C1999167 |
| Males | C0086582 |
| Chronic kidney disease | C1561643, C4075517, C4553188, C4075526 |
| COPD | C0024117 |
| Lung cancer | C0684249, C0242379, C1306460 |
| Immunocompromised state (weakened immune system) from solid organ transplant | C0029216, C0524930 |
| Obesity | C0028754, C1963185 |
| Serious heart conditions, such as heart failure, coronary artery disease, or cardiomyopathies | C0018802, C4554158, C0018801, C0010054, C1956346, C0878544, C0796094, C0020542 |
| Sickle cell disease | C0002895 |
| Asthma | C0004096, C2984299 |
| Neurologic conditions, such as dementia | C0002395, C0011265, C0497327, C0014544, C0026769, C0455388, C1417325, C0030567, C0036572 |
| Cerebrovascular disease (affects blood vessels and blood supply to the brain) such as stroke | C0678234, C1961121, C0549207, C1261287, C1522213, C0524466, C0038454, C0007282, C0595850, C0158570, C0002940 |
| Cystic fibrosis | C0010674 |
| Hypertension | C0020538, C1963138 |
| Immunocompromised state (weakened immune system) from blood or bone marrow transplant, immune deficiencies, HIV, use of corticosteroids, or use of other immune weakening medicines | C0005961, C3540726, C3540727, C0021051, C0279026, C3539185, C3540725, C0001617, C1955133 |
| Pregnancy | C0032961 |
| Liver diseases | C0023895 |
| Pulmonary fibrosis (having damaged or scarred lung tissues) | C4553408, C0034069 |
| Smoking | C1881674, C1548578, C0037369, C0453996 |
| Diabetes | C0011847, C0011849 |
| Thalassemia | C0039730, C0002312 |

### Analysis of Eligibility Criteria

#### Quantitative criteria

We used the open-source Valx tool [30] to extract and standardize the quantitative eligibility criteria from the COVID-19 studies. Valx is a system that can extract numeric expressions from free-text eligibility criteria and standardize them into a structured format. For example, from the inclusion criterion “BMI > 25 kg/m^2^”, the variable name “BMI”, the comparison operator “>“, the threshold value “25”, and the measurement unit “kg/m^2^” were extracted into 4 discrete fields. Valx is also able to recognize synonyms of a variable and convert the units to standard ones. We then analyzed the frequency of the quantitative criteria and the threshold values used for patient eligibility determination.

#### Qualitative eligibility features

To extract the qualitative eligibility features from COVID-19 studies, we used a new eligibility criteria parsing tool [29], which consists of a context-free grammar (CFG) and an information extraction (IE) modules to transform free-text eligibility criteria to structured formats for downstream analysis. The CFG module uses a lexer to divide criteria into tokens and a modified Cocke-Younger-Kasami algorithm to build parse trees from tokens, which are subsequently analyzed by removing duplicates and subtrees. The IE module uses an attention-based bidirectional long short-term memory with a conditional random field layer for named entity recognition to extract MeSH terms from criteria text. Based on the evaluation in [29], its performance is competitive in entity recognition, entity linking, and attribute linking. As it only extracted MeSH concepts from eligibility criteria but not their temporal constraints and other qualifiers, we called them “eligibility features” in this paper. To evaluate its concept extraction accuracy, we manually reviewed a random sample of 300 rows of extracted results along with their original criteria. In the extracted results, there are cases where a term was identified but was not matched to a MeSH concept. In such cases, we considered them to be false negatives. The precision is 98.9%. The recall is 81.1%. The false negative ones were mostly quantitative criteria (29.3%) or due to missing concepts in MeSH (48.3%). We manually corrected the parsing errors of the frequent concepts. For example, we corrected the parsing results of the criterion “men”, which was parsed as “multiple endocrine neoplasia”. It is fine to miss some quantitative criteria as they were extracted by Valx [30] with a high sensitivity and specificity. We also merged similar concepts in the parsing results based on the analysis needs. Detailed information about the merging of extracted concepts can be found in the Supplementary Material II. To evaluate the accuracy of extracting known risk factors from eligibility criteria using the tool [29], we reviewed a random sample of 300 rows of extracted risk factors, the precision is 100%. Regarding the recall, we took a random sample of 200 unique criteria. 72 of them contain a risk factor and the program extracted 61 of them, making the recall to be 84.72%. After the qualitative eligibility features of COVID-19 studies were parsed, we conducted three types of analyses: (1) frequency of the qualitative eligibility features; (2) clustering analysis of the clinical studies based on the parsed eligibility features; and (3) frequency of exclusion eligibility features on chronic conditions and risk factors. Since (1) is intuitive, we explain the process of (2) and (3) in details as follows.

#### Clustering analysis of clinical studies

We used the clustering analysis to group the clinical studies based on their eligibility features. After the inclusion and exclusion criteria are parsed by the aforementioned tool [29], we utilized the parsed concepts as features to construct clinical study representation. For inclusion and exclusion eligibility features, we first removed the duplicated concepts for each clinical study. For example, if “pregnancy women” is mentioned multiple times in the exclusion criteria, only one was kept. Then, we append the prefixes ‘inc’ or ‘exc’ to the concepts extracted from inclusion or exclusion criteria respectively to differentiate them. After data preprocessing, we constructed the data representations by treating each clinical study as a text document that contains concepts from inclusion and exclusion eligibility features. The Term Frequency-Inverse Document Frequency (TF-IDF) weighting scheme was employed to construct the feature vectors to feed to the K-means clustering algorithm [31]. K-means is rather easy to implement and apply on large and high dimensional data sets. The algorithm assigns the instance to one of the clusters. The objective is to minimize the sum of the distances of the instances within the cluster to the cluster centroid. The silhouette value and CHindex were jointly used to measure the clustering results of K-means to determine the optimal number of clusters. The silhouette values measures similarity of an instance to its own cluster compared to other clusters. In this research, we experimented with k values from 2 to 50 for k-means. The optimal k was chosen when the silhouette value average of all instances is high and there are at least 20 instances for each cluster. To visualize the clustering result, we employed the uniform manifold approximation and projection (UMAP) [32] to project the high dimensional data into two-dimensional space for visualization. The UMAP reduces the dimensions by estimating the topology of the high dimensional data. It considers the local relationships within groups and global relationships between groups. UMAP can be applied directly to sparse matrices. In addition, we also clustered the interventional studies considering both the extracted eligibility features and the enrollment values. The reason we included only interventional studies in this analysis is that observational studies often have a huge enrollment value, thereby dominating the clustering results. In addition, the eligibility criteria of observational studies are usually broad and unrestrictive. We used Principle Component Analysis (PCA) to reduce the dimensionality of eligibility features (weighted by TF-IDF) to 10 and then added two more features: the enrollment value (normalized to 0-1) and the intervention type, due to their importance deemed by the study team. We used UMAP to visualize the clustering result following the same process as the one using only eligibility features.

#### Exclusion eligibility features on chronic conditions and risk factors

First, we examined the upper limit and lower limit of the age criterion, which are structured data in the study summaries. Then, from the results of the criteria parsing tool [29], we examined the use of exclusion eligibility features about 15 most prevalent chronic conditions among older adults in the National Inpatient Sample of the Healthcare Cost and Utilization Project (HCUP) (appearing in over 6% of the older adults in NIS) [33]. These conditions include hypertension, hyperlipidemia, ischemic heart disease, diabetes, anemia, chronic kidney disease, atrial fibrillation, heart failure, chronic obstructive pulmonary disease and bronchiectasis, rheumatoid arthritis or osteoarthritis, acquired hypothyroidism, Alzheimer disease and related disorders or senile dementia, depression, osteoporosis, and asthma. In addition, we also considered three chronic conditions that are prevalent in all adults: cancer, stroke, and high cholesterol. We then analyzed the use of risk factors that may lead to serious illnesses in the eligibility criteria.

All the data and codes pertaining to this project have been deposited to GitHub: https://github.com/ctgatecci/Covid19-clinical-trials.

## Results

### Basic Characteristics of the COVID-19 Clinical Studies

Table 2 shows the basic characteristics of 3,765 COVID-19 clinical studies in ClinicalTrials.gov. Among 3,765 clinical studies included in this paper, a majority of them are interventional studies (clinical trials). Among those interventional studies, 43.4% are in Phase 2 or 3. Most of the studies (83.19%) are sponsored by hospitals, universities, research institutes, or individuals. Besides drugs, other interventions include biological (15.9%), behaviors (7.06%), device (6.01%), diagnostic test (3.57%), and others (10.72%, e.g., genetic, dietary supplements, radiation, and combination). The majority of the studies focused on treatment (40.88%) and prevention (8.95%). The 10 most frequently tested drugs in different clinical studies are Hydroxychloroquine (N=162), Azithromycin (N=56), Tocilizumab (N=36), Ivermectin (N=31), Favipiravir (N=28), Remdesivir (N=28), Ritonavir (N=21), Lopinavir (N=20), Interferon (N=20), Plasma (N=19) (Figure 1).

**Table 2.**
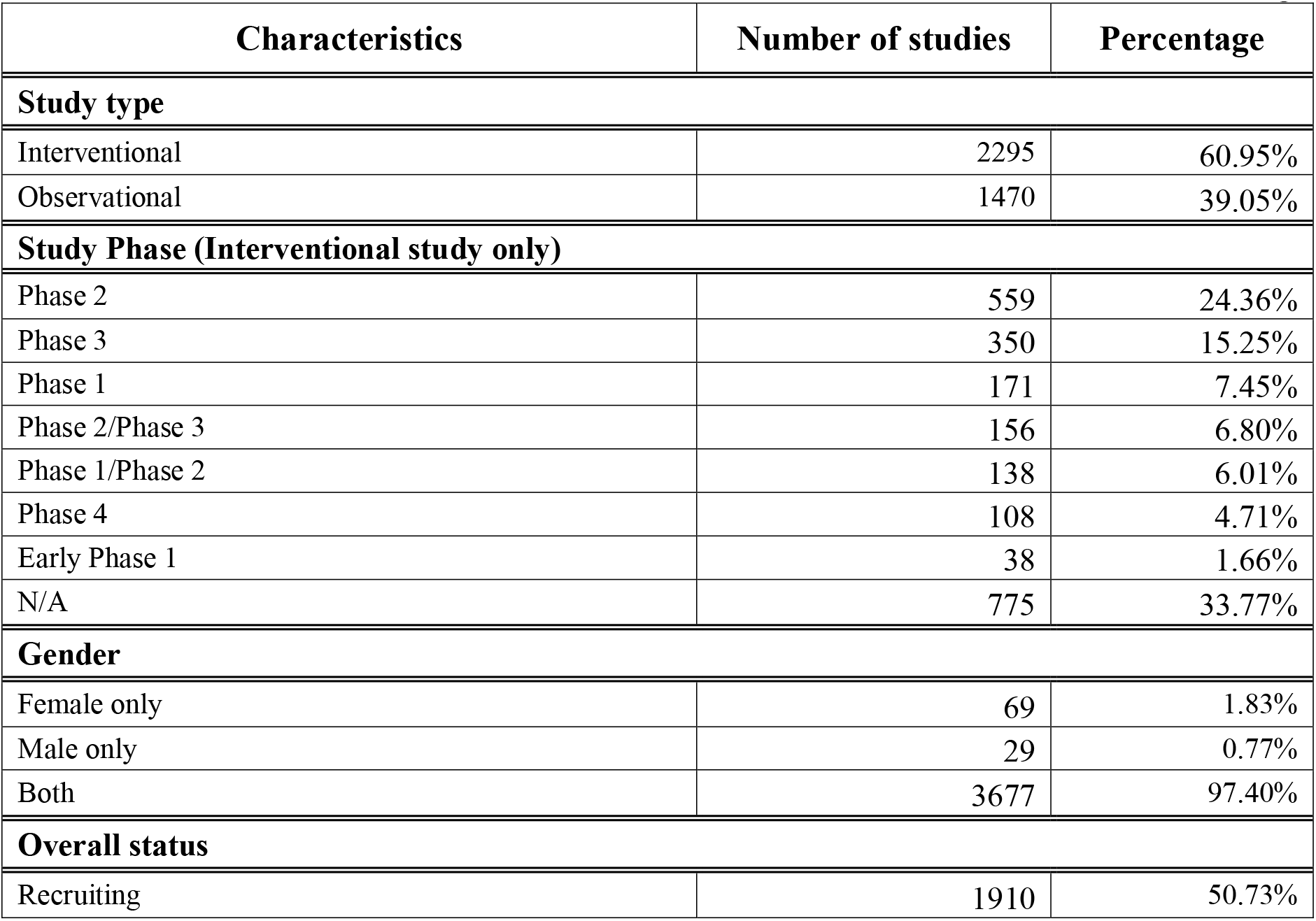
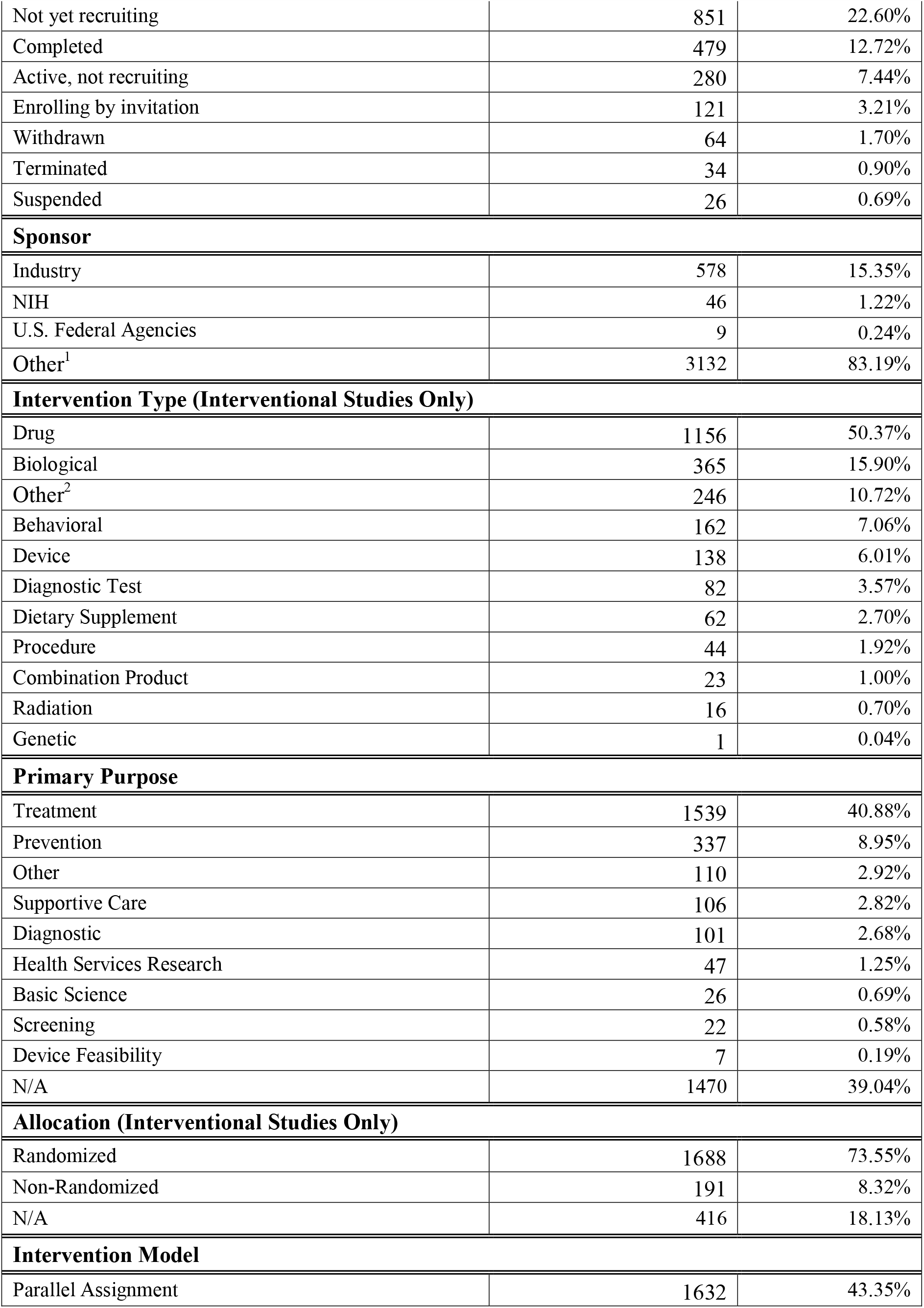
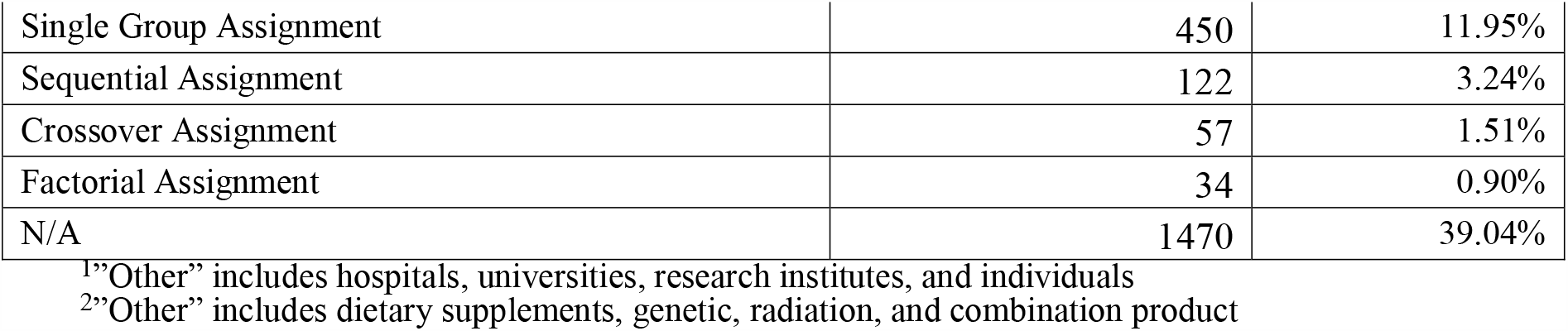
Basic characteristics of 3,765 COVID-19 clinical studies in ClinicalTrials.gov.

**Figure 1.**
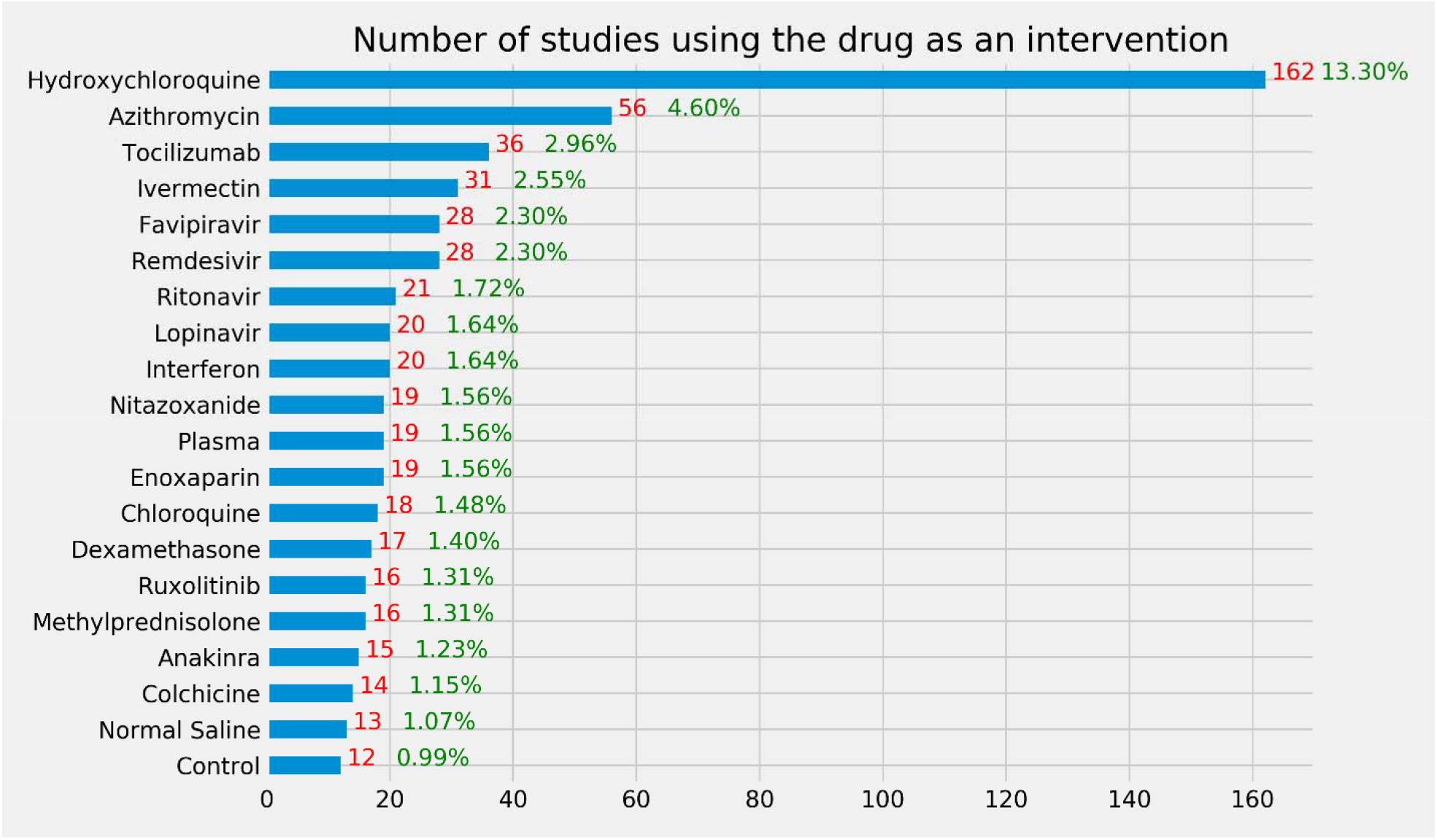
Number of interventional studies using a drug as an intervention. The denominator is the 1218 interventional studies using drug as an intervention. Note that some studies tested multiple drugs.

### Risk Factors in Trial Description

Figure 2 illustrates the occurrences of the risk factors in the study description of the included studies. We merged the brief summary and detailed description. “Weak immune 1” corresponds to immunocompromised state from solid organ transplant and “weak immune 2” corresponds to immunocompromised state from blood or bone marrow transplant, immune deficiencies, HIV, use of corticosteroids, or use of other immune-weakening medicines. The top 5 risk factors mentioned in trial description are diabetes, hypertension, weak immune 2, obesity, and pregnancy. According to the t-test result, on average, interventional studies mentioned fewer risk factors in trial description than observational studies (mean value: 0.2 vs 0.27, *P* = 0.002, two-tailed t-test). The number of risk factors mentioned in trial description was significantly associated with the intervention type (p < 0.001, ANOVA), while no statistically significant association between it and the primary purpose (P = 0.078, ANOVA).

**Figure 2.**
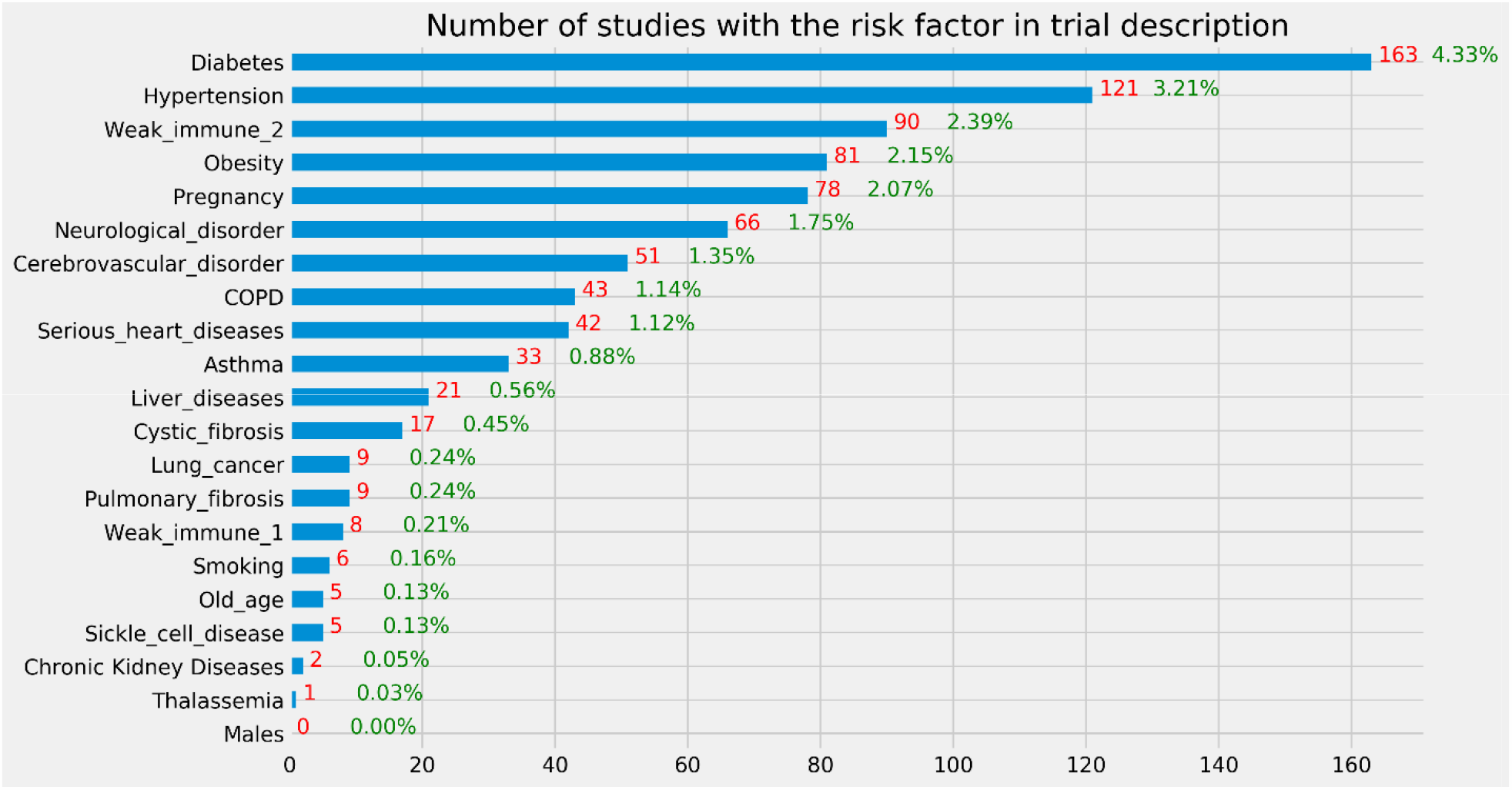
Number of studies with a risk factor for severe illness in the trial description. The denominator is the 3765 clinical studies included in this study.

### Quantitative criteria

Table 3 lists the top 20 frequently used quantitative criteria in COVID-19 clinical studies. Note that the “age” criterion is also a structured field in the study records. Based on the analysis of upper age limit, 67.3% (N= 2534) clinical studies do not have an upper age limit. For those that have an upper age limit, the most frequent limits are 80 (N=191), 75 (N=117), 65 (N=108), 100 (N=99), and 70 (N=94). Regarding the lower age limit, only 9.8% studies (N=369) do not have lower age limit. Most frequently used lower age limits are 18 (N=2856), 16 (N=59), 20 (N=49), 19 (N=32), and 50 (N=31). Figure 3 illustrates the percentage of COVID-19 clinical studies that consider each age range. In general, patients who are over 18 years old are considered while those over 70 years old are less considered than 18-70 years old. Regarding oxygen saturation, most studies use 93% (N=114), 94% (N=58), or 90% (N=29) as threshold values.

**Table 3.** Top 20 frequently used quantitative criteria in COVID-19 clinical studies.

| Rank | Criteria | Frequency | Percentage | Rank | Criteria | Frequency | Percentage |
| --- | --- | --- | --- | --- | --- | --- | --- |
| 1 | Age | 2255 | 70.4% | 11 | Platelet count | 108 | 3.4% |
| 2 | Oxygen saturation | 229 | 7.2% | 12 | ANC | 106 | 3.3% |
| 3 | Pao2/fio2 | 223 | 7.0% | 13 | Creatinine clearance | 105 | 3.3% |
| 4 | BMI | 222 | 6.9% | 14 | Heart rate | 75 | 2.3% |
| 5 | Respiratory rate | 190 | 6.0% | 15 | Diastolic blood pressure | 64 | 2.0% |
| 6 | AST | 185 | 5.8% | 16 | QTC | 63 | 2.0% |
| 7 | EGFR | 145 | 4.5% | 17 | Total bilirubin level | 56 | 1.8% |
| 8 | Temperature | 131 | 4.1% | 18 | Pulse rate | 52 | 1.6% |
| 9 | Systolic blood pressure | 113 | 3.5% | 19 | Hemoglobin | 48 | 1.5% |
| 10 | ALT | 109 | 3.4% | 20 | Creatinine | 46 | 1.4% |

**Figure 3.**
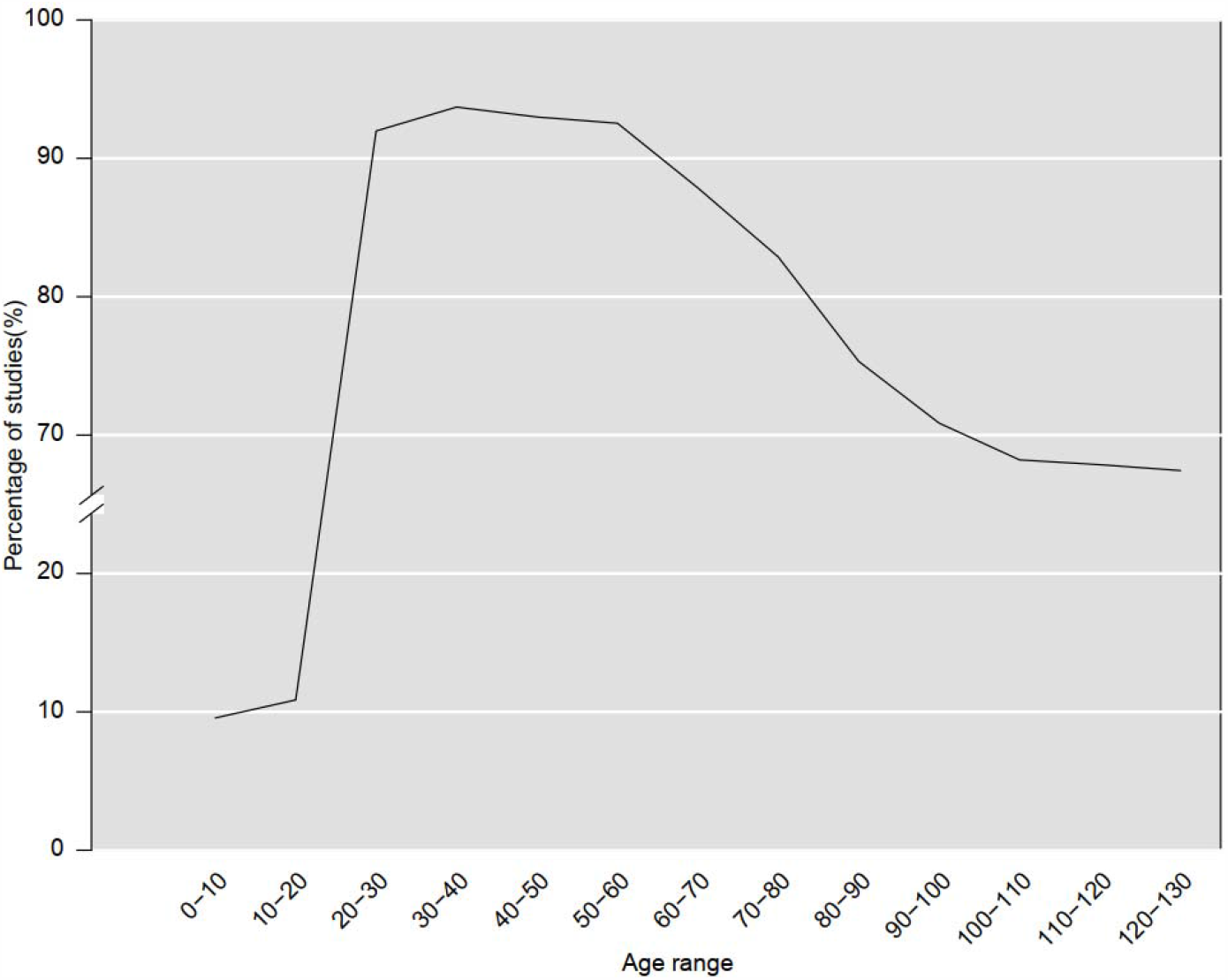
Percentage of COVID-19 clinical studies allowing age ranges

### Qualitative Eligibility Features

Figure 4 illustrates frequent concepts extracted from inclusion and exclusion criteria of COVID-19 clinical studies. According to these results, COVID-19 diagnosis, polymerase chain reaction, pneumonia, diabetes, therapeutics, mechanical ventilation were often used eligibility features in the inclusion criteria, whereas pregnancy, therapeutics, kidney diseases, cancer, HIV, mechanical ventilation, hydroxychloroquine, hepatitis C were often used eligibility features in the exclusion criteria.

**Figure 4.**
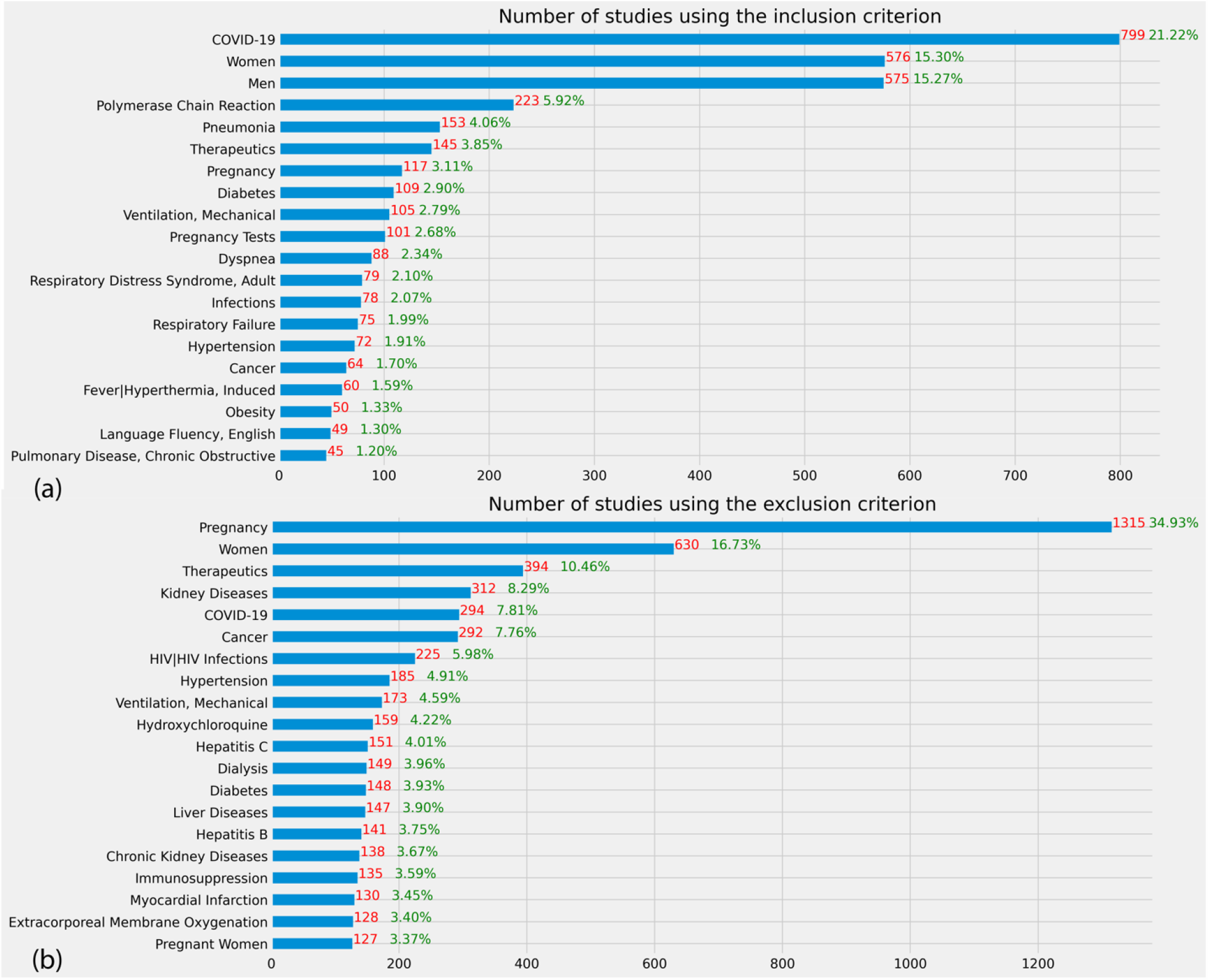
Frequent eligibility features of COVID-19 clinical studies. The denominator is the 3765 clinical studies included in this study.

Figure 5 shows the number of studies that used an exclusion eligibility feature about a common chronic condition prevalent among older adults in the included studies. Even though a majority of studies did not exclude patients with these chronic conditions, some highly prevalent chronic conditions such as cancer, heart failure, hypertension, and chronic kidney disease, COPD, and diabetes are among the most frequently used exclusion criteria in 2.76% - 8.42% studies. Few studies purposely included patients with a risk factor that may lead to serious illnesses, but few studies explicitly excluded them except for pregnant women. According to the results of the statistical tests, on average, interventional studies used more risk factors in eligibility criteria than observational studies (mean: 1.19 vs. 0.22, p<0.001, two-tailed t-test). There is a statistically significant association between the number of risk factors used in eligibility criteria and the intervention type (p < 0.001, ANOVA), and primary purpose of the studies (P< 0.001, ANOVA).

**Figure 5.**
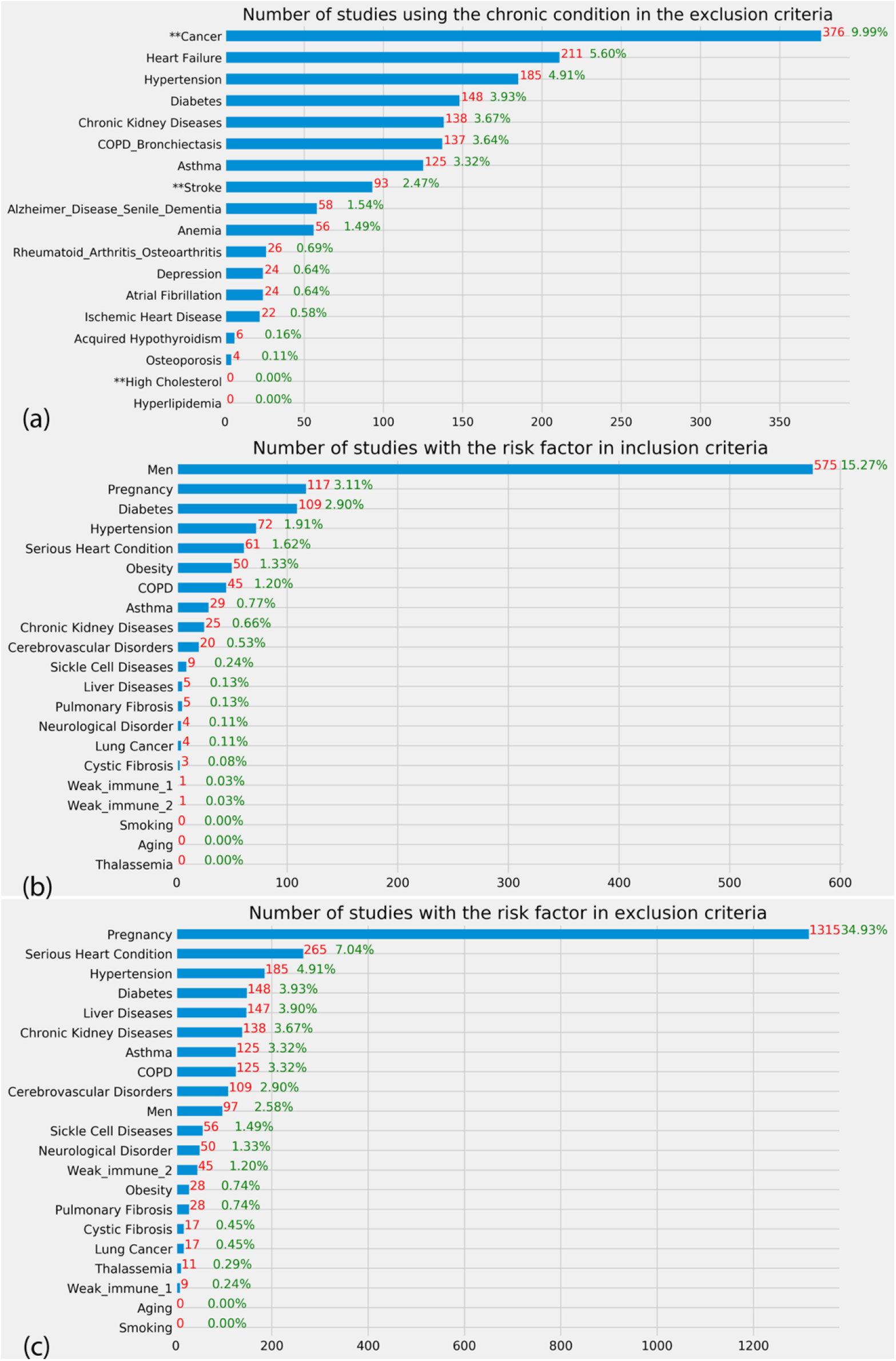
(a) Number of studies using a prevalent chronic condition among the older adults in exclusion criteria. ** represents the conditions that are not in the list of top 15 prevalent conditions among older adults but prevalent in younger adults. (b) Number of studies with the risk factor in inclusion criteria (c) Number of studies with the risk factor in exclusion criteria. The denominator of these three figures is the 3765 clinical studies included in this study.

Table 4 shows the top 10 frequent inclusion and exclusion features used in the studies in each of the 7 clusters resulting from the clustering analysis with eligibility features only. Pregnancy is the most frequent exclusion criterion in all the 7 clusters. Studies in Cluster #0 often included patients with pneumonia and excluded patients with cognition/cognitive behavioral therapy/cognitive dysfunction. Studies in Cluster #1 often excluded patients who are on therapeutics, kidney diseases, and cancer. Studies in Cluster #2 often included patients with polymerase chain reaction. Studies in Cluster #5 often excluded HIV/HIV Infections, Hepatitis B, Hepatitis C, and Cancer. Figure 6 shows the visualization of these 7 clusters using UMAP. The detailed results of the clustering analysis of the COVID-19 clinical studies are provided in the Supplementary Material III. The results of the clustering analysis of the interventional studies when considering the eligibility features, the enrollment, and the intervention type are provided in the Supplementary Material IV (table and figure) and V (detailed results).

**Table 4.**
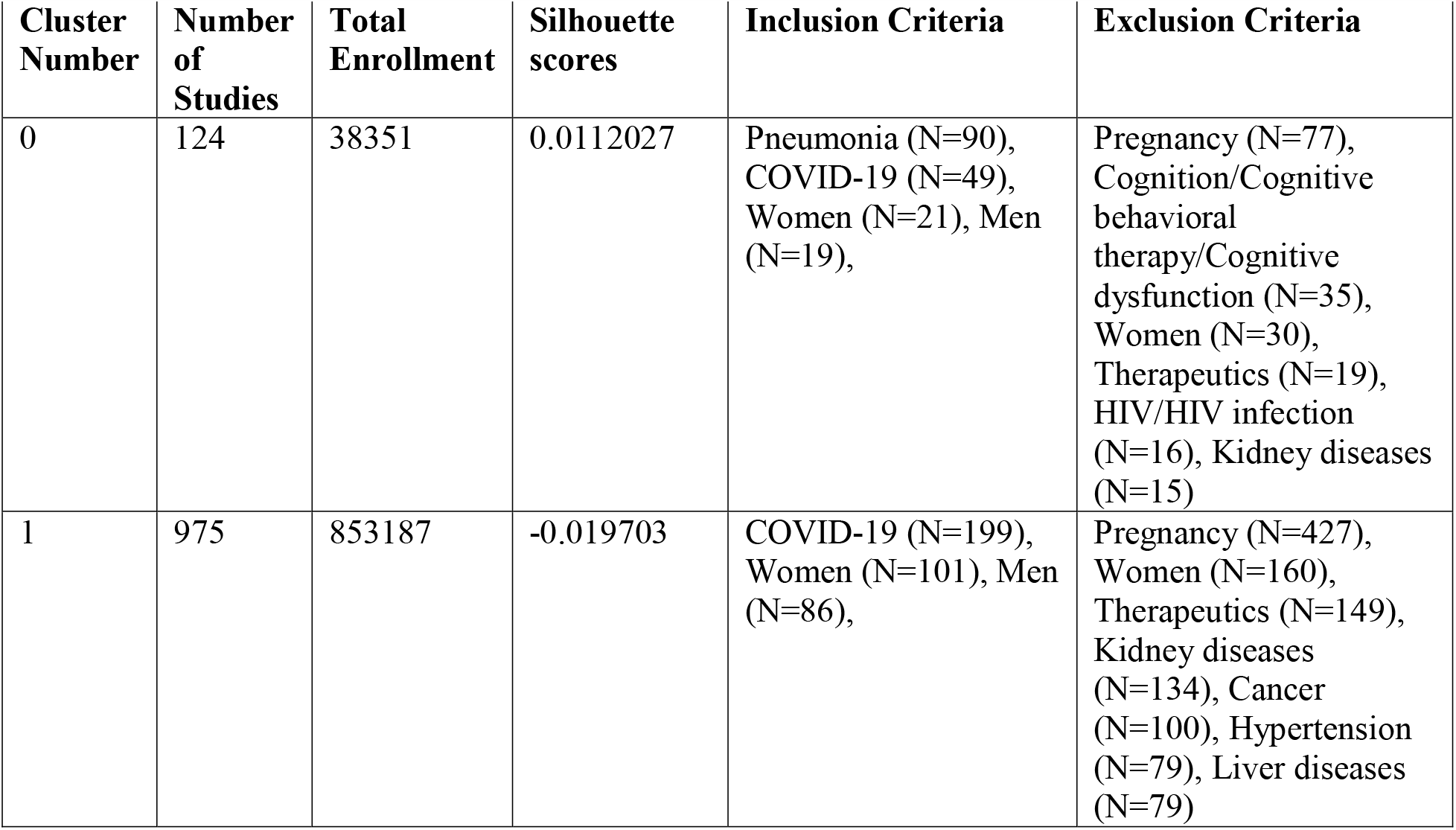
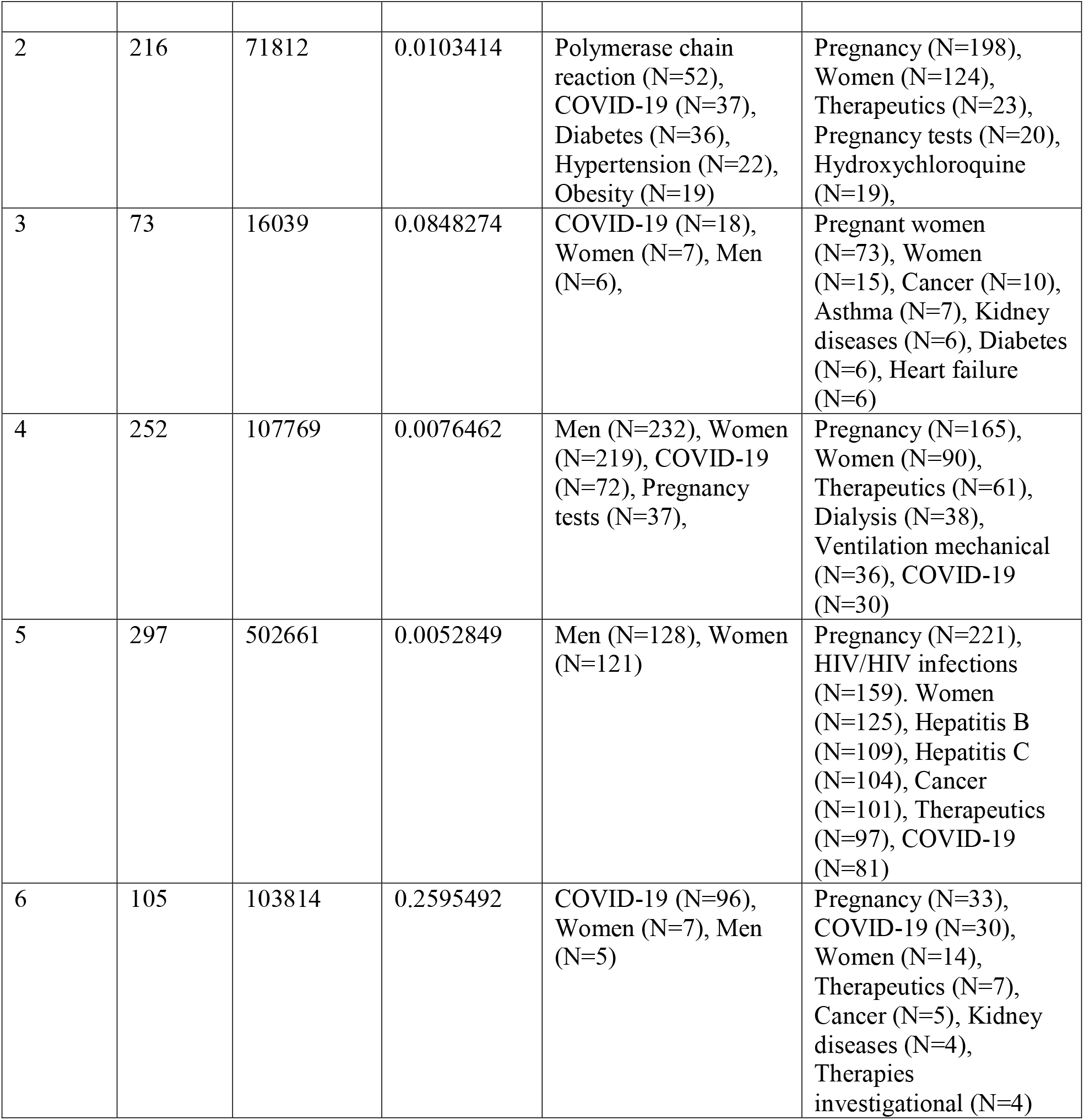
Top 10 frequently used concepts in inclusion criteria and exclusion criteria of the studies in each cluster of the clustering analysis with eligibility features.

**Figure 6.**
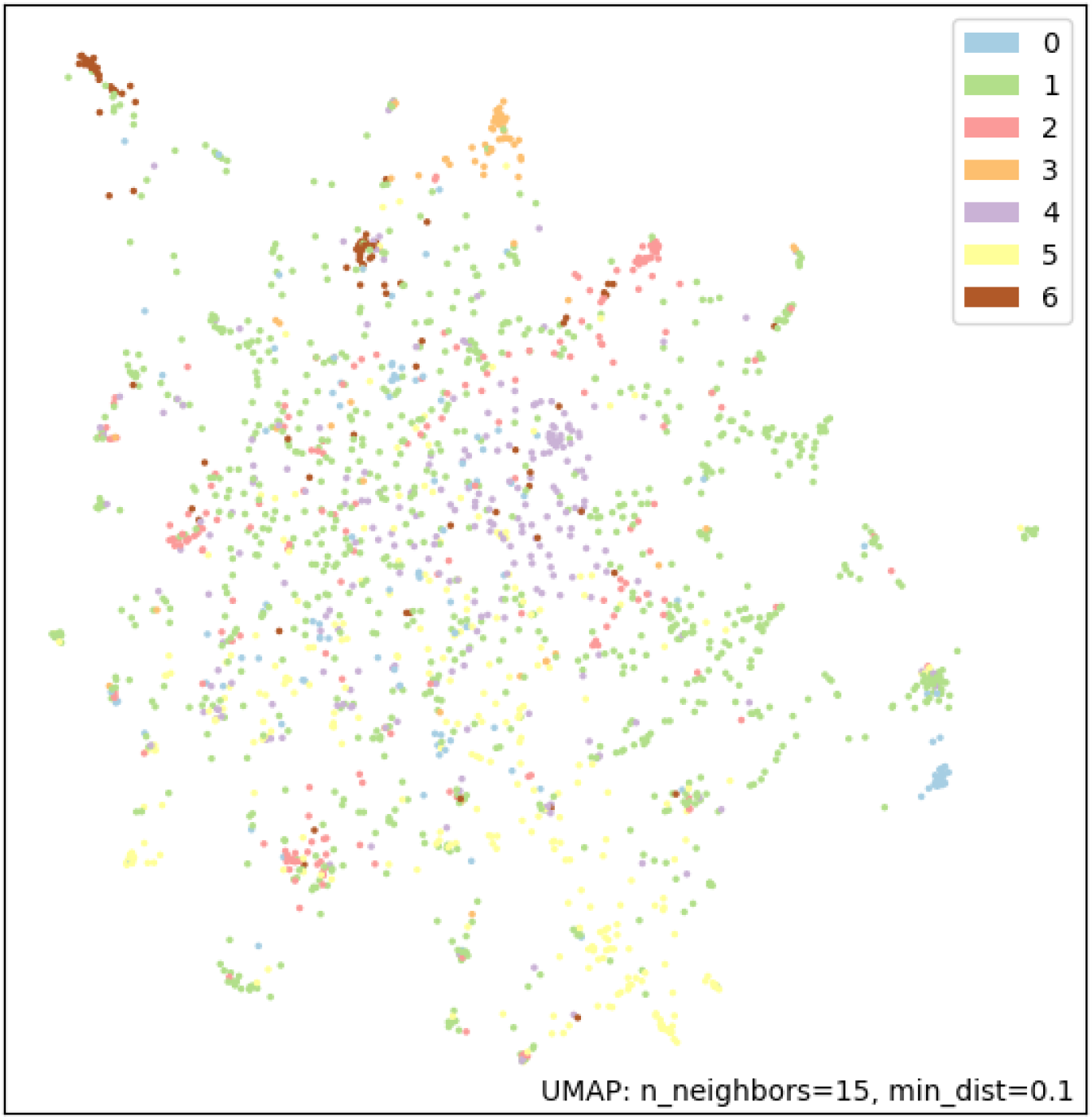
Visualization of the 7 clusters using UMAP

## Discussion

As the novel coronavirus COVID-19 has significantly impacted our lives and even taken lives of hundreds of thousands of people in the past 8 months, we must quickly identify repurposed drugs or develop new drugs and vaccines to safely and effectively control the spread of the virus and save lives. Clinical studies, especially randomized controlled trials, are a fundamental tool used to evaluate the efficacy and safety of new medical interventions for disease prevention or treatment. Many clinical studies are being conducted to find safe and effective treatments and vaccines. Thus far, significant efforts have been devoted to repurposing existing FDA-approved drugs including immunosuppression (e.g., Hydroxychloroquine, Tocilizumab), anti-virus (e.g., Fevipiravir, Lopinavir/Ritonavir), anti-parasite (e.g., Ivermectin, Nitazoxanide), antibiotics (e.g., Azithromycin), and anticoagulant (e.g., Enoxaparin). In our analysis of the COVID-19 clinical studies, we found that the use of eligibility criteria and consideration of risk factors in these studies did not change much from June 18, 2020 to November 27, 2020 even though the number of COVID-19 studies in ClinicalTrials.gov grew from 2,192 to 4,028.

To transform clinical trials and lower their cost, a notion of “digital clinical trial” was created to leverage digital technology to improve important aspects such as patient access, engagement, and trial measurement [34]. The US National Institutes of Health and the National Science Foundation held a workshop in April 2019 about the implementation of digital technologies in clinical trials, in which “defining and outlining the composition and elements of digital trials” and “elucidating digital analytics and data science approaches” were identified as two of the five top priorities. As COVID-19 is a major health crisis that impacts people regardless of their age, gender, and race/ethnicity, it is in our interest to understand if clinical studies on COVID-19 adequately considered the representation of real-world populations. Based on our analysis, most clinical studies consider both genders (97.4%, N=3,667), do not have an upper age limit 67.3% (N= 2,534), and have a lower age limit of 18 (75.9%, N=2,856). The exclusion of children in these studies may be due to lower susceptibility and lower rates of mortality and hospitalization for children with COVID-19 compared to adults [35]. As serious illnesses of COVID-19 mostly occurred in older adults with underlying health conditions, it is not surprising that they are in general considered by most COVID-19 studies, based on our analysis of their eligibility criteria. Most studies did not set an upper age limit (67.3%, N= 2534) and did not exclude older adults with common chronic conditions. This is contrary to the recent New York Times articles conjecturing that older people are left out form COVID-19 trials [10]. As older adults are the most likely to be hospitalized due to COVID-19, clinicians may be more likely to choose to include them to fulfill the sample size requirement of the trials. Nonetheless, conducting COVID-19 clinical studies could still be challenging in the traditional clinical trial eco-system, where patient accrual is often delayed due to logistical constraints [36]. The generalizability of the study results to the real-world population should be evaluated with state-of-the-art techniques [9]. Older adults could have still been underrepresented in COVID-19 clinical studies due to logistical reasons, which can only be assessed with the published results after the completion of the studies [37]. In addition, pregnant women are often excluded in COVID-19 studies. Even though pregnant women are in general excluded in most clinical trials due to the potential risks to both the women and the unborn babies, observational studies should carefully evaluate the vertical transmission of the virus and negative impact of COVID-19 on the well-being of mothers and infants [38]. Clinical studies should adequately evaluate the efficacy and safety of treatments and vaccines on vulnerable population groups.

### Limitations

A few limitations should be noted. First, some data in ClinicalTrials.gov are missing. For example, 33.8% (N=775) of the interventional studies miss study phase information. 39% (N=1,470) of studies do not have primary purpose information. Second, we relied on the search function of ClinicalTrials.gov when retrieving COVID-19 studies. There may be study indexing errors, but the scale should be minimal and would not impact the findings. Third, we used the QuickUMLS and the new eligibility criteria parsing tool [29] to extract risk factors, chronic conditions, disorders, and procedures from study records. Thus, the sensitivity and specificity of the term extraction and normalization are dependent on the quality of the UMLS Metathesaurus and the eligibility criteria parsing tool. Nonetheless, we have carefully curated the term extraction results to ensure that our results are as accurate as possible.

## Conclusions and Future Work

In this paper, we systematically analyzed COVID-19 clinical study summaries in ClinicalTrials.gov using natural language processing. Specifically, we analyzed whether these clinical studies considered the underlying health conditions (and other risk factors) that may increase the severity of the COVID-19 illness. Given the ongoing nature of this pandemic, it is inevitable that early trials will start with different knowledge of risk factors than later trials. In future work, we will perform a longitudinal analysis of COVID-19 studies to assess the changes in the use of eligibility criteria and consideration of risk factors for severe illness in COVID-19 patients. As results of COVID-19 studies become available, we will be able to assess the extent to which the trial design and eligibility criteria in particular would impact the findings as well as the real-world population representativeness of these studies using generalizability assessment methods [9].

## Data Availability

All the data and codes pertaining to this project have been deposited to GitHub.

https://github.com/ctgatecci/Covid19-clinical-trials

## Acknowledgments

We would like to sincerely thank Markku Salkola for his generous help with the eligibility criteria parsing tool.

## Data Availability

The data underlying this article were accessed from ClinicalTrials.gov. The derived data generated in this research will be shared on reasonable request to the corresponding author.

## Author Contribution

ZH conceived, designed, guided, and coordinated the study and the writing. ZH collected the data from ClinicalTrials.gov, performed the data analyses, interpreted the results, and drafted the manuscript. AE performed the natural language processing of the clinical study records. XL performed the clustering analysis. AX performed statistical tests to assess the association between the occurrences of risk factors and the study characteristics. All the authors edited the manuscript thoroughly. The submitted manuscript has been approved by all the authors.

## Footnotes

1 We did not evaluate the recall due to lack of a benchmark dataset for trial description with all the risk factors annotated manually.

