## Supplementary material for "How the clinical research community responded to the COVID-19 pandemic: An analysis of the COVID-19 clinical studies in ClinicalTrials.gov": Supplementary_Material_II.pdf

**Part 1. Number of studies with inclusion and exclusion criteria**

1. We dropped duplicated rows across ‘nct_id’, ’eligibility_criteria’, and ‘concepts’ columns in the parsing results, since we were interested in number of studies using a certain inclusion criterion concept or a certain exclusion criterion concepts. By dropping duplicated rows, we made sure that the same clinical studies will not be counted more than once.
2. After evaluating the Facebook Clinical Trial Parser tool, we found that some concepts should be merged. Here is a list of combined concepts:
    - Changed 'Men|Multiple Endocrine Neoplasia Type 2a|Multiple Endocrine Neoplasia Type 2b’ to 'Men'; Since ‘MEN’ is the abbreviation for the ‘Multiple Endocrine Neoplasia’ and the parser misidentified ‘Men’ as ‘Multiple Endocrine Neoplasia’;
    - Combined 'Immunosuppression|Immunosuppression (Physiology)' with 'Immunosuppression' as 'Immunosuppression';
    - Combined 'Pregnancy, Unplanned', 'Pregnancy, Multiple' , and 'Pregnancy' as 'Pregnancy';
    - Combined ‘Diabetes Mellitus’, ’Diabetes Mellitus, Type 1|Diabetes Mellitus, Type 2’, ’ Diabetes Mellitus, Type 1’, ’Diabetes Mellitus, Type 2’, ’Diabetes Insipidus’ as ‘Diabetes’;
    - Combined ‘Hypertension, Pulmonary’, ‘Hypertension, Portal’, ‘Intracranial Hypertension’, ‘Familial Primary Pulmonary Hypertension|Idiopathic Pulmonary Arterial Hypertension|Pulmonary Hypertension, Primary’, ‘Hypertension, Pregnancy-Induced’,and ‘Hypertension, Malignant’ as ‘Hypertension’;
    - Combined ‘Liver Diseases’ and ‘Liver Diseases, Alcoholic’ as ‘Liver Diseases’;
    - Combined ‘Obesity|Obesity Management|Obesity, Morbid’ and ‘Obesity’ as ‘Obesity’;
    - Combined ‘Renal Insufficiency, Chronic', 'Chronic Kidney Diseases’ and ‘Chronic Kidney Diseases’ as ‘Chronic Kidney Diseases’;
    - Combined ‘Kidney’, ‘Kidney Failure’, ‘Kidney Failure, Chronic’, ‘Kidney Diseases’, and ‘Kidney Failure, Acute’ as ‘Kidney Diseases’;
3. After combining some concepts, we showed the number of studies in inclusion concepts and exclusion concepts.

**Part 2. Number of studies with the risk factor in exclusion criteria and inclusion criteria.**

Step 1 is the same as Step 1 in Part 1

We have a list of risk factors. For each risk factor, we have combined similar MeSH concepts in the parsing results. After combining, we showed the number of studies with the risk factors in exclusion concepts and inclusion concepts. Here is the list of combined concepts:

- Changed 'Men|Multiple Endocrine Neoplasia Type 2a|Multiple Endocrine Neoplasia Type 2b’ to 'Men'; Since ‘MEN’ is the abbreviation for the ‘Multiple Endocrine Neoplasia’, the parser misidentified ‘Men’ as ‘Multiple Endocrine Neoplasia’;
- Combined 'Pregnancy, Unplanned', 'Pregnancy, Multiple' , and 'Pregnancy' as 'Pregnancy';
- Combined ‘Diabetes Mellitus’, ’Diabetes Mellitus, Type 1|Diabetes Mellitus, Type 2’, ’ Diabetes Mellitus, Type 1’, ’Diabetes Mellitus, Type 2’, ’Diabetes Insipidus’ as ‘Diabetes’;

- Combined ‘Hypertension, Pulmonary’, ‘Hypertension, Portal’,  ‘Intracranial Hypertension’, ‘Familial Primary Pulmonary Hypertension|Idiopathic Pulmonary Arterial Hypertension|Pulmonary Hypertension, Primary’, ‘Hypertension, Pregnancy-Induced’,and ‘Hypertension, Malignant’  as ‘Hypertension’;
- Combined ‘Liver Diseases’ and ‘Liver Diseases, Alcoholic’ as ‘Liver Diseases’;
- Combined ‘Renal Insufficiency, Chronic','Chronic Kidney Diseases’ and ‘Chronic Kidney Diseases’ as ‘Chronic Kidney Diseases’;
- Combined ‘Obesity|Obesity Management|Obesity, Morbid’ and ‘Obesity’ as ‘Obesity’;
- Combined ‘Heart Failure’, ‘Congestive Heart Failure’, ‘Heart Failure, Systolic’, ‘Coronary Artery Disease’, and Cardiomyopathies’ as ‘Serious Heart Condition’;
- Combined ‘Cerebrovascular Disorders’, ‘Stroke’, ‘Stroke, Acute’ as ‘Cerebrovascular Disorder’;
- Changed ‘Bone Marrow Transplantation’ to ‘Weak_immune_2’;
- Combined ‘Anemia’, ‘Anemia, Sickle Cell’, ‘Anemia, Aplastic’, ‘Anemia, Hemolytic’, and ‘Fanconi Anemia’ as ‘Sickle Cell Diseases’;
- Combined ‘Asthma, Exercise-Induced’ and ‘Asthma’ as ‘Asthma’;
- Changed ‘Organ Transplantation’ to ‘Weak_immune_1’;
- Changed ‘Dementia’ to ‘Neurological Disorder’;
- Combined ‘beta-Thalassemia’, ‘Thalassemia’, and ‘alpha-Thalassemia’ as ‘Thalassemia’;
- Changed 'Pulmonary Disease, Chronic Obstructive' to ‘COPD’;
- Combined ‘Pulmonary Fibrosis’ and ‘Idiopathic Pulmonary Fibrosis’ as ‘Pulmonary Fibrosis’.

**Part 3. Number of studies using the chronic condition in exclusion criteria**

Step 1 is the same as Step 1 in Part 1.

- We have a list of common chronic condition. For each risk factor, we combined similar MeSH concepts in the parsing results. After combining, we showed the number of studies with the chronic condition concepts in exclusion criteria. Here is a list of the combined concepts:
- Combined ‘Hypertension, Pulmonary’, ‘Hypertension, Portal’,  ‘Intracranial Hypertension’, ‘Familial Primary Pulmonary Hypertension|Idiopathic Pulmonary Arterial Hypertension|Pulmonary Hypertension, Primary’, ‘Hypertension, Pregnancy-Induced’,and ‘Hypertension, Malignant’  as ‘Hypertension’;
- Changed ‘'Myocardial Ischemia' to 'Ischemic Heart Disease’;
- 'Combined ‘Diabetes Mellitus’, ’Diabetes Mellitus, Type 1|Diabetes Mellitus, Type 2’, ’ Diabetes Mellitus, Type 1’, ’Diabetes Mellitus, Type 2’, ’Diabetes Insipidus’ as ‘Diabetes;
- Combined ‘Anemia’, ‘Anemia, Sickle Cell’, ‘Anemia, Aplastic’, ‘Anemia, Hemolytic’, and ‘Fanconi Anemia’ as ‘Anemia’;
- Combined ‘Renal Insufficiency, Chronic','Chronic Kidney Diseases’ and ‘Chronic Kidney Diseases’ as ‘Chronic Kidney Diseases’;
- Combined ‘Heart Failure’, ‘Congestive Heart Failure’, and  ‘Heart Failure, Systolic’ as ‘Heart Failure’;

- Combined 'Pulmonary Disease, Chronic Obstructive' and 'Bronchiectasis' as 'COPD_Bronchiectasis';
- Changed 'Arthritis, Rheumatoid' to 'Rheumatoid_Arthritis_Osteoarthritis;
- Changed 'Hypothyroidism' to 'Acquired Hypothyroidism';
- Combined 'Alzheimer Disease' and 'Dementia' as 'Alzheimer_Disease_Senile_Dementia'
- Combined 'Asthma' and 'Asthma, Exercise-Induced' as 'Asthma';
- Combined ‘Cancer’, ‘Lymphoma’, ‘Prostate Cancer’, ‘Breast Cancer’, ‘Cancer of skin’, ‘Multiple Myeloma’, ‘Cancer of the Uterine Cervix’, ‘Basal Cell Cancer’, ‘Cancer of Colon’, ‘Lymphoma, Large-Cell, Anaplastic’, ‘Cancer|Carcinoma in Situ’, ‘Lymphoma, Mantle-Cell’, ‘Urinary Bladder Cancer’, ‘Melanoma’, and ‘Lymphoma, Follicular|Lymphoma, Follicular, Grade 3’ as Cancer;
- Combined ‘Stroke’ and ‘Stroke, Acute’ as ‘Stroke’.
