## Supplementary material for "How the clinical research community responded to the COVID-19 pandemic: An analysis of the COVID-19 clinical studies in ClinicalTrials.gov": Supplementary_Material_IV.pdf

**Appendix Table 1.** Top 10 frequently used concepts in inclusion criteria and exclusion criteria of the studies in each cluster of the clustering analysis with eligibility features, intervention type, and enrollment.

| Cluster Number | Number of Studies | Total Enrollment | Silhouette scores | Intervention Type | Inclusion Criteria | Exclusion Criteria |
|---|---|---|---|---|---|---|
| 0 | 120 | 96834 | 0.1652518 | Drug (117), Dietary (2), Supplement (2), Other (1) | COVID-19 (36), Men (27), Women (27) | Hydroxychloroquine (114), Pregnancy (79), Chloroquine (79), Women (46), Therapeutics (33), Glucosephosphate dehydrogenase deficiency (32), Azithromycin (31) |
| 1 | 532 | 667023 | 0.4460913 | Biological (329), Behavioral (95), Device (87), Product (21), Combination (21) | Women (139), Men (134), COVID-19 (113), | Pregnancy (259), Women (133), Cancer (94), Therapeutics (92), COVID-19 (85), HIV/HIV infections (77), Hypertension (67) |
| 2 | 135 | 52626 | 0.006419 | Drug (100), Other (13), Diagnostic (9), Test (9), Dietary (5), Supplement (5), Device (4), Procedure (2), Radiation (1), Behavioral (1) | Men (130), Women (119), COVID-19 (22), | Pregnancy (83), Women (41), Therapeutics (25), Cancer (15), Kidney diseases (15), COVID-19 (14), Dialysis (13) |
| 3 | 23 | 12864 | 0.3585205 | Drug (11), Other (5), Diagnostic (2), Test (2), Biological (2), Device (1), Procedure (1), Dietary (1), Supplement (1) | Polymerase chain reaction (6) | Pregnancy (23), Women (12), COVID-19 (1) |
| 4 | 833 | 694010 | 0.3420431 | Drug (611), Other (129), Dietary (35), Supplement (35), Procedure (24), Diagnostic (23), Test (23), Radiation (11) | Women (137), Men (123), Polymerase chain reaction (78) | Pregnancy (480), Women (225), Therapeutics (146), Kidney diseases (130), Cancer (103), Ventilation mechanical (82), HIV/HIV infections (77) |
| 5 | 52 | 32242 | 0.2909869 | Drug (13), Biological (10), Device (9), | COVID-19 (52), Women (3), Men (3), | Pregnancy (15), Women (6), COVID-19 (4), Cancer (1) |

| | | | | | | |
|---|---|---|---|---|---|---|
| | | | | Diagnostic (7), Test (7). Other (7), Behavioral (3), Procedure (2), Dietary (1), Supplement (1) | Polymerase chain reaction (1), Respiratory failure (1), Diabetes (1) | |
| 6 | 33 | 9378 | 0.2354992 | Drug (8), Device (8), Behavioral (8), Other (7), Biological (1), Dietary (1). Supplement (1) | COVID-19 (8), Women (4) | Cognition/Cognitive behavioral therapy/Cognitive dysfunction (33), Pregnancy (13), Hypertension (7), Women (5), Therapeutics (5), Kidney diseases (4), Cancer (4), Immunosuppresive agents (3) |
| 7 | 49 | 8874 | 0.0995203 | Drug (16), Other (8), Diagnostic (6), Test (6), Device (6), Biological (6), Dietary (5), Supplement (5), Procedure (2) | COVID-19 (9), Men (5), Women (5), Pneumonia (3), Polymerase chain reaction (3) | Pregnant women (49), Women (9), Cancer (4), Asthma (4), Diabetes (3) |
| 8 | 265 | 119782 | 0.1314706 | Drug (215), Other (26), Dietary (8), Supplement (8), Procedure (8), Radiation (4), Diagnostic (3), Test (3), Device (1) | COVID-19 (256), Men (57), Women (53), Pneumonia (27) | Pregnancy (169), Women (81), Therapeutics (55), COVID-19 (40), Kidney diseases (35), Ventilation mechanical (34) |

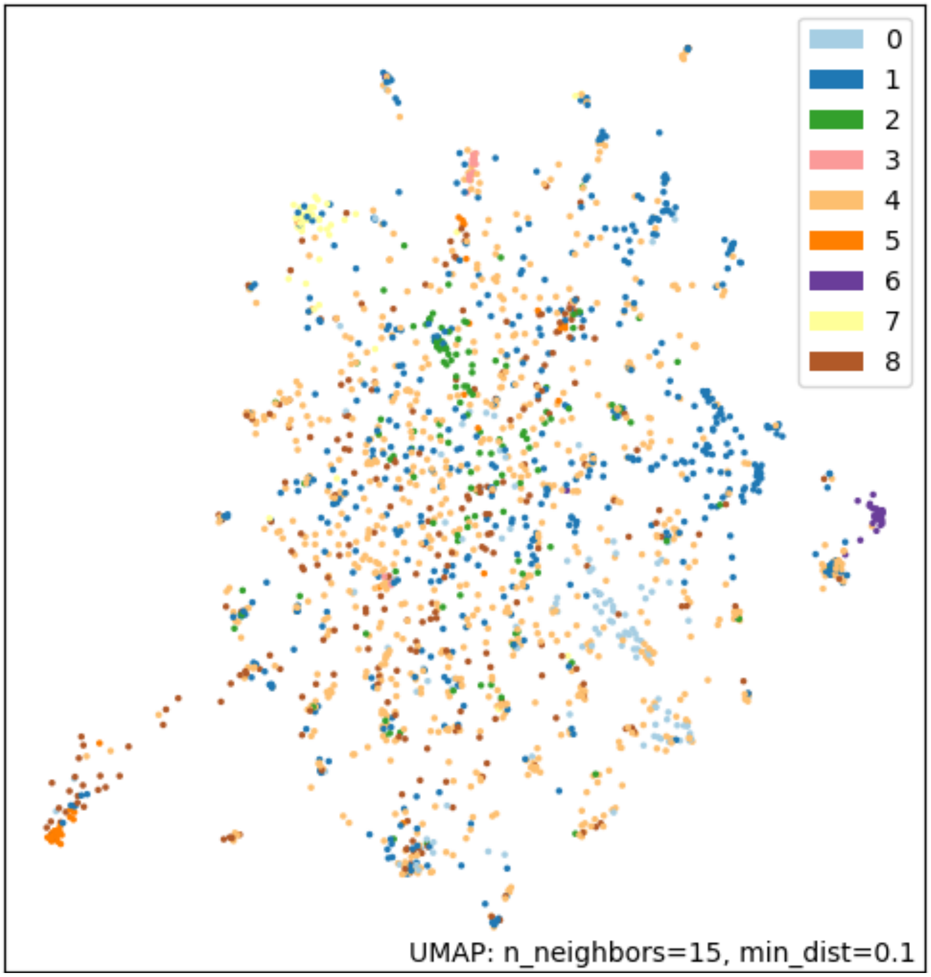


**Appendix Figure 1.** Visualization of the 9 clusters using UMAP
